# Epigenetic aging and perceived psychological stress in old age

**DOI:** 10.1101/2022.02.24.22271447

**Authors:** Valentin Max Vetter, Johanna Drewelies, Yasmine Sommerer, Christian Humberto Kalies, Vera Regitz-Zagrosek, Lars Bertram, Denis Gerstorf, Ilja Demuth

## Abstract

Adverse effects of psychological stress on physical and mental health, especially in older age, are well documented. How perceived stress relates to the epigenetic clock measure, DNA methylation age acceleration (DNAmAA), is less well understood and existing studies reported inconsistent results.

DNAmAA was estimated from five epigenetic clocks (7-CpG, Horvath’s, Hannum’s, PhenoAge and GrimAge DNAmAA). Cohen’s Perceived Stress Scale (PSS) was used as marker of psychological stress.

We analyzed data from 1,100 Berlin Aging Study II (BASE-II) participants assessed as part of the GendAge study (mean age = 75.6 years, SD = 3.8 years, 52.1% women).

In a first step, we replicated well-established associations of perceived stress with morbidity, frailty, and symptoms of depression in the BASE-II cohort studied here. In a second step, we did not find any statistically significant association of perceived stress with any of the five epigenetic clocks in multiple linear regression analyses that adjusted for covariates.

Although the body of literature suggests an association between higher DNAmAA and stress or trauma during early childhood, the current study found no evidence for an association of perception of stress with DNAmAA in older people. We discuss possible reasons for the lack of associations and highlight directions for future research.

## Introduction

Greater overall psychological stress can have adverse effects on health and is associated with higher mortality [1]. Its association with cardiovascular disease [2], upper respiratory disease [3], symptom severity of rheumatoid arthritis [4], depressive symptoms [5] and other phenotypes [6, 7] is well documented. Several pathways have been proposed to link psychological stress with morbidity. First, psychological stress has been shown to result in poor health decisions and promote impulsive decision-making [8] such as increased consumption of nicotine or alcohol [9], other substance abuse [10] and sleep deprivation leading to an increased risk for numerous diseases [11]. Second, two major endocrine response pathways mediate the physiological response to psychological stress. First, catecholamines released by the sympathetic-adrenal-medullary (SAM) system affect the cardiovascular, the pulmonary, and the immune system and prepare the body to fight or flee if threatened [12]. Second, the hypothalamic-pituitary-adrenocortical axis (HPA) regulates the level of glucocorticoids which have immunosuppressive and anti-inflammatory effects and promote gluconeogenesis [13, 14]. Both systems, if activated repeatedly and for long durations, are known to increase the risk for disease [15-17]. This is partially mediated via down-regulation of glucocorticoid receptors and a chronic state of inflammation [18].

Furthermore, the reactivity of the HPA axis to psychological stress increases with age [19]. Despite poorer physiological regulation in the face of stressors in older age, older adults might have advantages in both the overall exposure as well as emotional response to stressors [20, 21]. For instance, older adults are able to evade stressful situations more successfully than younger adults by using secondary coping or avoidance strategies [21, 22]. Importantly, however, in situations where older adults are confronted with an unavoidable stressor, these emotional advantages may become unfavorable in the face of physiological vulnerability, e.g., a disabling disease [21, 23].

How psychological stress impacts physical and mental health depends on numerous modifiable and non-modifiable factors [6]. Especially poorer health seems to increase vulnerability to stress-induced disease in older age [6]. One way of objectifying age-dependent biological vulnerability is through the measurement of biomarkers of aging. One promising biomarker in this domain is the determination of DNA methylation age (DNAm age) and, in particular, its deviation from chronological age, DNAm age acceleration (DNAmAA) [24]. Both markers are estimated from epigenetic clocks that utilize data on the methylated fraction of specific cytosin-phosphat-guanine (CpG) sites. Several such epigenetic clocks are available which differ in the way they were designed and which aspects of aging they represent best [25]. Previous work has suggested that an association between psychological stress and DNAm age appears plausible due to the fact that 85 of the 353 CpG sites of the Horvath clock (and possibly CpG subfractions of other DNAm clocks as well) are located within glucocorticoid response elements (GRE) [26, 27]. These DNA sequences represent binding sites to glucocorticoid receptors and were shown to be at or near to CpG sites that were especially affected by glucocorticoid dependent demethylation mediated by demethylating enzymes and decreased expression of DNA methyltransferase [28, 29]. Furthermore, the number of CpG sites within GRE’s exceeds the amount that would be expected by chance [29].

Additionally, epigenetic changes were suggested to be a possible link [30, 31] between adverse childhood experiences and mortality as well as higher morbidity burden in late life [32]. It was proposed that this link could be mediated by health-adverse coping mechanisms (activated as a result of high levels of anxiety and depression) that are associated with adverse childhood experiences [33]. Some of these coping strategies, such as smoking, alcohol abuse and and a high BMI resulting from unhealthy eating habits, were shown to be associated with DNAmAA in some studies [34-36]. However, these results were not unequivocally replicated [37-39] (reviewed in ref. [40]).

Previous studies that examined the relationship between DNAmAA and stress operationalized stress as low socioeconomic status (SES) [41, 42], (childhood) trauma [26, 43-45], racial discrimination [46], or exposure to violence [47]. Many previous studies on the topic focused on changes in DNAm age during childhood as this period is known to be particularly prone to stress-related epigenetic changes [29].

In contrast, in this work we focus on older age which was shown to be the second most vulnerable phase in a person’s life in terms of epigenetics [29]. As epigenetic modifications remain even after the psychological stimulus has ceased there is the possibility of cumulating effects on the epigenome exerted by repeated psychological stressors [29]. Specifically, we analyzed the association between the amount of experienced stress (measured by Cohen’s Perceived Stress Scale [PSS] [48]) and several DNAm age estimators (i.e. the 7-CpG clock [49], Horvath’s clock [50], Hannum’s clock [51], PhenoAge [34], GrimAge [52]) and in 1,100 older adults. While the PSS represents a well-established marker of perceived stress [48], to our knowledge it has not been investigated in the context of epigenetic aging before. While we were able to replicate well-established associations with perceived stress, none of the five epigenetic clocks analyzed in the current study were associated with the perception of stress.

## Methods

### BASE-II/GendAge Study

BASE-II is a longitudinal study that aims to identify factors that promote healthy aging. Participants were recruited through advertisements in local newspapers and on public transport in the greater Berlin area, Germany. At baseline examination (2009-2014), 2,171 participants were medically examined (∼75% aged 60–84 years and ∼25% aged 20–37 years; this latter, younger group was not considered in the present work). In this study, we focus on the cross-sectional analysis of 1,083 BASE-II participants of the older age group who were reexamined on average 7.4 years after baseline as part of the GendAge study. Seventeen additional BASE-II participants were available for follow-up that were not included in the medical baseline examination. For a more detailed cohort information at baseline and follow-up, please refer to Bertram et al. [53], Gerstorf et al. [54], and Demuth et al. [55].

All participants gave written informed consent. The medical assessments at baseline and follow-up were conducted in accordance with the Declaration of Helsinki and approved by the Ethics Committee of the Charité – Universitätsmedizin Berlin (approval numbers EA2/029/09 and EA2/144/16). They were registered in the German Clinical Trials Registry as DRKS00009277 and DRKS00016157.

### Measures

#### Perceived Stress

Stress was assessed by eight items of the Perceived Stress Scale (PSS) that was developed by Cohen, Kamarck and Mermelstein in 1983 [48]. Participants answered the questions on a scale from 1 (“never”) to 5 (“very often”). The answers were averaged and z-transformed with R’s “scale” function for the linear regression analyses. Data on PSS was available for 1,006 participants of the GendAge study.

### DNA methylation age (DNAm age)

DNAm age was estimated by five epigenetic clocks. The 7-CpG clock was developed from methylation data obtained through methylation-sensitive single nucleotide primer extension (MS-SNuPE) from samples collected at baseline examination of the participants analyzed in this study [49] and replicated in a separate cohort [56]. To estimate DNAm age using the 7-CpG clock, bisulfite converted DNA samples were amplified with multiplex PCR. Subsequently, the PCR products were cleaned and underwent MS-SNuPE. Finally, the luminescent signals of the SNuPE products were measured on a “3730 DNA Analyzer” (Applied Biosystems, HITACHI) [57, 58] and the methylation fraction was calculated as peak height ratio. For a more detailed description of the methods used see ref. [59].

Additionally, DNAm age was estimated using Horvath’s clock [50], Hannum’s clock [51], PhenoAge [34] and GrimAge [52] from methylation data determined with the “Infinium MethylationEPIC” array (Illumina, Inc., USA). Briefly, probes were filtered according to the detection *p*-value. Probes with more than 1% of samples having a detection *p*-value of 0.05 were removed from the analysis, as well as probes with a bead count smaller than 3 in more than 5% of the samples. Outliers were identified with the *outlyx* function and the *pcout* function with a threshold of 0.15 [60]. Additionally, samples with a bisulfite conversion efficiency below 80% (as estimated by the *bscon* function) were removed. Subsequently, the samples were reloaded with outliers excluded and normalized with the function *dasen*. The function *qual* was used to determine the extent of change in beta values in each sample due to normalization. Samples with a root-mean-square deviation of 0.1 or larger were removed and loading and normalization were repeated with the new sample set with removed outliers. Finally, non-cg probes, probes affected by SNPs [61], probes aligning to multiple locations in the genome [62], as well as probes on the X and Y chromosome were removed. On the EPIC array only 512 of the original 513 CpG sites and 64 of the original 71 CpG sites were available for the estimation of PhenoAge and Hannum’s clock, respectively. For more information on the protocol used to obtain DNAm data at follow-up, see ref. [59].

### DNAm age acceleration (DNAmAA)

To correct for the well documented association between blood celltype composition and chronological age, we employed a blood cell count adjusted model to calculate DNAmAA [37, 63]. It was calculated as unstandardized residuals of a linear regression analysis of DNAm age on chronological age and leukocyte cell distribution (neutrophils, monocytes, lymphocytes, and eosinophils in G/l). Blood cell composition was measured by an accredited clinical biochemistry laboratory (MVZ Labor 28 GmbH, Berlin, Germany) using automated standard methods (flow cytometry).

### Morbidity, depressive symptoms, and frailty measure

Morbidity burden was assessed using a modified version [64] of Charlson’s morbidity index [65]. Symptoms of depression were recorded with the Center for Epidemiological Studies Depression Scale (CES-D) [66]. A score of 16 or more points on the 0 to 60 point scale is used to identify individuals at risk for clinical depression [67] but the full scale was used in this study to make use of more subtle differences in depressive symptoms as well. Frailty was measured using Fried’s frailty phenotype [68] that incorporates unintentional weight loss, self-reported exhaustion, weakness (grip strength), slow walking speed (timed-up-and-go test), and low physical activity [69].

### Covariates

We included the following covariates in all statistical models to account for potential confounding: Differences between sexes with respect to aging [70] and DNAmAA [38, 71], and the effect of psychological stress on disease [13] are well documented. Therefore, sex was included as covariate in all regression analyses. In addition, we performed sex-stratified analyses for all tests. Other covariates included were information on alcohol consumption (“yes”/”no”) and smoking behavior (packyears) which were assessed in one-to-one interviews by trained study personnel. The body mass index (BMI) was calculated using electronic height and weight measurements (via a “seca 763” measuring station, SECA, GERMANY). Educational attainment was assessed as education years until highest degree [72]; this information was available for 994 participants. Lastly, we controlled for genetic ancestry by using the first four principal components from a principal component analysis on genome-wide SNP genotyping data [73] generated in the same individuals.

### Statistical analyses

All statistical analyses were executed in R 3.6.2 [74]. Linear regression analyses were performed using the “lm” function, and all figures were produced with the “ggplot2” package [75].

Participants were only excluded from an analysis if they were missing a variable required for the respective analysis (available case analysis). We indicate the number of observations for each analysis individually. A *p*-value below 0.05 was considered statistically significant.

## Results

### Sample characteristics

Cross-sectional data on 1,100 participants were available. Included participants were between 64.9 and 94.1 years old (mean age: 75.6 years, SD = 3.8 years, 52.1% female). Perception of stress, assessed as averaged answer on eight items of Cohen’s Perceived Stress Scale (PSS), was normally distributed (Figure 1) and no sex-difference was found (*t-test*, p=0.08, Supplementary Table 1). Men had statistically significant higher DNAmAA in all five available epigenetic clocks (*t-test, p* ≤0.001, Supplementary Table 1). This sex-difference was reported before in this data set [76] as well as in others [38, 71].

**Figure 1:**
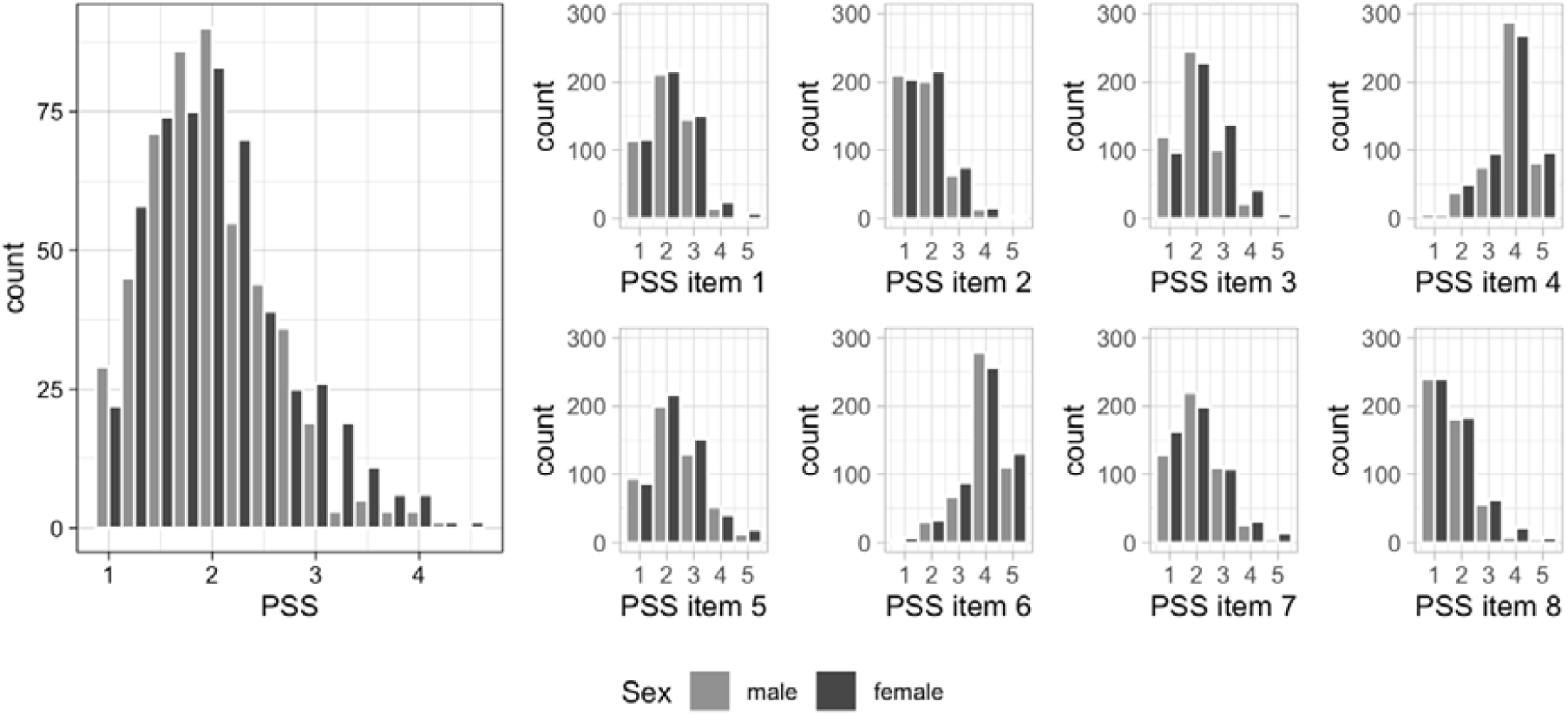
Distribution of Cohen’s PSS in GendAge participants (n=1,006). Please note that items four and six were reversed before inclusion in the final PSS. Note: PSS = Perceived Stress Scale.

### Association between Cohen’s PSS and depressive symptoms, morbidity, and frailty

In a first step, we tested for known associations between psychological stress and clinical phenotypes. To improve the interpretability of the PSS, it was z-transformed prior to the inclusion in linear regression models. The known associations between stress and depressive symptoms, morbidity and frailty were also observed in this data set (Table 2). Specifically, an increase of one standard deviation on the PSS was associated with 0.3 points higher morbidity index, 0.2 points higher frailty score, and 0.8 points higher scores on the CES-D, after adjustment for all covariates. These associations persisted in sex-stratified analyses and seemed to be particularly pronounced in women, as the morbidity index and the CES-D showed a higher effect size in this subgroup compared to men (Supplementary Table 2).

**Table 1:** Cohort characteristics of 1,100 BASE-II participants of the GendAge study.

|  | n | % | mean | sd | min | max |
| --- | --- | --- | --- | --- | --- | --- |
| Chronological age (years) | 1100 |  | 75.60 | 3.77 | 64.91 | 94.07 |
| Sex (female) | 573 | 52.09 |  |  |  |  |
| Smoking (packyears) | 1019 |  | 9.79 | 17.61 | 0.00 | 150.00 |
| BMI | 1098 |  | 26.97 | 4.25 | 17.17 | 49.68 |
| Education (years) | 994 |  | 14.42 | 2.92 | 7 | 18 |
| Alcohol intake (yes) | 912 | 83.14 |  |  |  |  |
| Frailty score | 1087 |  | 0.76 | 0.87 | 0.00 | 4.00 |
| CES-D | 1089 |  | 13.55 | 3.71 | 0.00 | 35.00 |
| Morbidity index | 954 |  | 1.39 | 1.54 | 0.00 | 9.00 |
| 7-CpG clock DNAmAA | 1071 |  | 0.03 | 6.42 | -24.93 | 34.48 |
| Horvath's clock DNAmAA | 1067 |  | 0.03 | 4.04 | -12.31 | 23.45 |
| Hannum's clock DNAmAA | 1067 |  | 0.01 | 3.89 | -10.80 | 28.57 |
| PhenoAge DNAmAA | 1067 |  | 0.04 | 5.42 | -16.54 | 25.80 |
| GrimAge DNAmAA | 1067 |  | 0.03 | 3.39 | -10.82 | 12.84 |
| PSS | 1006 |  | 2.08 | 0.64 | 1.00 | 4.50 |

Note: BMI = body mass index, CES-D = Center for Epidemiologic Studies Depression Scale, DNAmAA = DNA methylation age acceleration, PSS = perceived stress scale.

**Table 2:** Multiple linear regression analyses of morbidity index, frailty score or CES-D on Cohen’s PSS in older BASE-II participants of the GendAge study. Linear regression models were adjusted for covariates. Model 1: no adjustment; Model 2: chronological age, sex; Model 3: Model 2 + smoking (packyears), alcohol (yes/no), BMI, and education.

| Dependent Variable | Model | Estimate | SE | *p*-value |  | n |
| --- | --- | --- | --- | --- | --- | --- |
| Morbidity Index | 1 | 0.222 | 0.052 | <0.001 | *** | 874 |
|  | 2 | 0.222 | 0.052 | <0.001 | *** | 874 |
|  | 3 | 0.259 | 0.055 | <0.001 | *** | 752 |
| Fried's Frailty Phenotype | 1 | 0.182 | 0.027 | <0.001 | *** | 995 |
|  | 2 | 0.178 | 0.027 | <0.001 | *** | 995 |
|  | 3 | 0.181 | 0.029 | <0.001 | *** | 852 |
| CES-D | 1 | 0.888 | 0.112 | <0.001 | *** | 997 |
|  | 2 | 0.873 | 0.112 | <0.001 | *** | 997 |
|  | 3 | 0.842 | 0.119 | <0.001 | *** | 854 |

Note: SE = standard error, CES-D = Center for Epidemiologic Studies Depression Scale.

### Relationship between DNAmAA and Cohen’s PSS

The potential relationship between Cohen’s PSS and the various DNAmAA parameters was assessed by multiple linear regression models. The fully adjusted model included sex, smoking, alcohol, BMI, education, and genetic ancestry as covariates (*Model 3*).

While weak associations were observed in the unadjusted (*Model 1*) and sex-adjusted model (*Model 2*) between PSS and Horvath’s DNAmAA (ß=-0.27, SE=0.13, p=0.04, n=976, *Model 2*) and PSS and PhenoAge DNAmAA (ß=-0.37, SE=0.17, p=0.04, n=976, *Model 2*), these were no longer significant in the full model (*Model 3*, p > 0.2, Table 3). Although no statistically significant association between PSS and 7-CpG, Hannum’s and GrimAge DNAmAA were found in unadjusted linear regression models, the coefficients pointed in the same negative direction. This was true for the sex-adjusted model (Model 2) as well, except for the relationship between PSS and GrimAge DNAmAA, were the beta-coefficient was slightly positive (ß= 0.013, p= 0.899, *Model 2)*. Like the results reported for the full dataset, sex-stratified subgroup analyses revealed no statistically significant associations after adjustment for covariates (Supplementary Table 3).

**Table 3:**
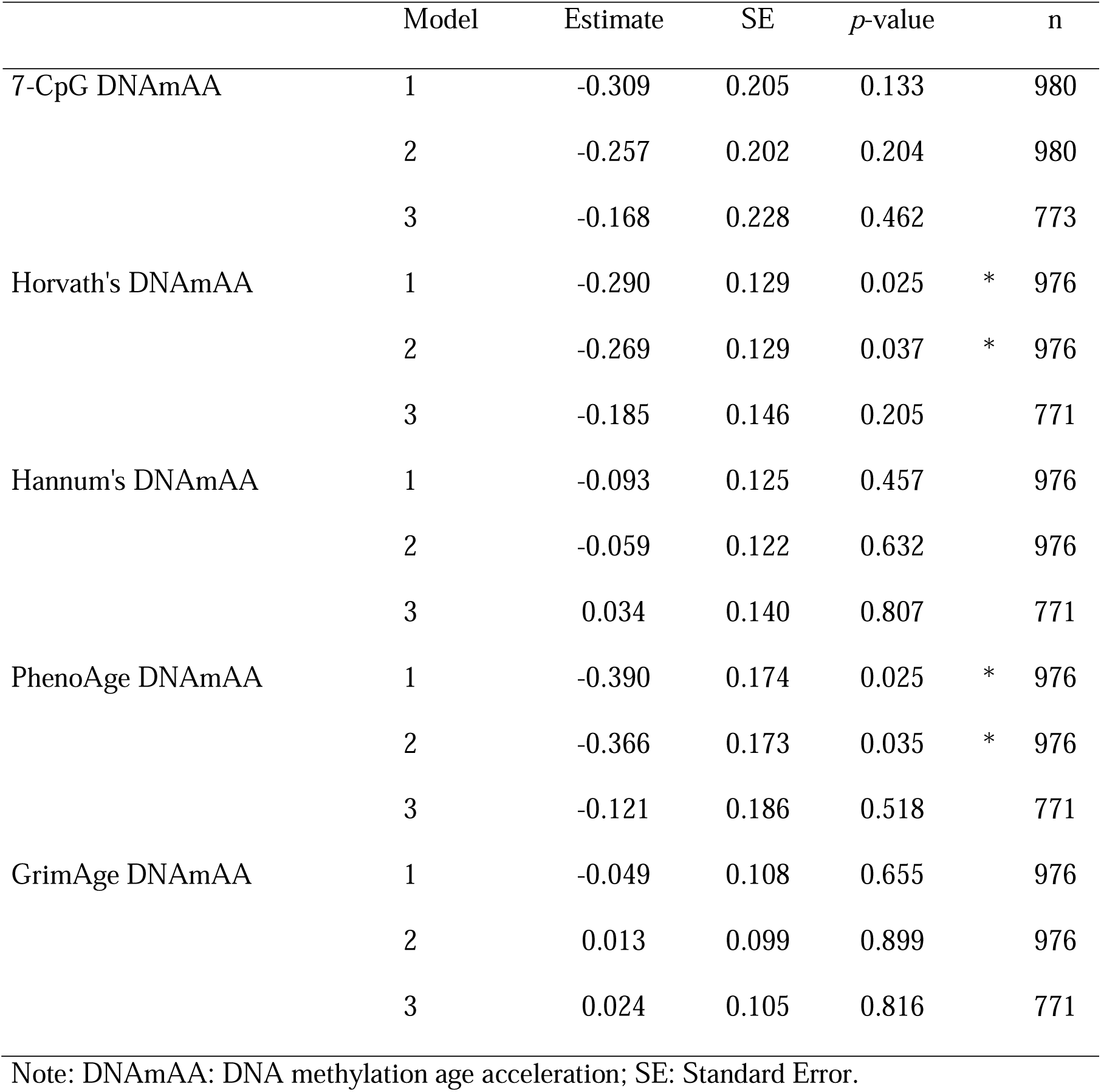
Multiple linear regression of Cohen’s PSS on DNAmAA of five epigentic clocks and covariates. Model 1: no covariates; Model 2: sex; Model 3: sex, smoking (packyears), alcohol intake (yes/no), BMI, education, and genetic ancestry.

## Discussion

In this study, we report data on perceived stress as assessed by the Cohen’s PSS and the biomarker DNAmAA as derived from five different epigenetic clocks in a comparatively large sample of older adults. Overall, we found no noteworthy associations between our marker of psychological stress and DNAmAA estimated by any of the five employed epigenetic clocks.

While our study is not the first on the topic, comparability with previous work is limited due to substantial differences in cohort characteristics and in quantification of stress as well as DNAmAA. Most previous studies focused on associations between retrospectively assessed life adversities during childhood and DNAmAA in comparatively young cohorts (with a mean age of 50 years or younger, reviewed in ref. [32]). A particular impact of psychological stress that was (retrospectively remembered as having been) experienced during childhood and adolescence on epigenetic changes was shown and explained by an high vulnerability to epigenetic changes during early age [29].

In contrast, only very few studies examined psychological stress during adulthood and its impact on epigenetic aging. In these studies, stress was most often operationalized as low socioeconomic status (SES) (overview in Supplementary Table 4). Similar to childhood and adolescence, older adults were reported to be especially prone to stress-related epigenetic changes, mostly due to a decline of the epigenetic maintenance system [29]. For instance, low income was associated with higher DNAmAA (using Hannum’s clock estimate) in a cohort of 100 black women (mean age 48.5 years) [39]. In a different study, Fiorito and colleagues found several measures for low socioeconomic status in a meta-analysis of three cohorts from Italy, Australia and Ireland (n=5,111, mean age: 57.28 years) to be associated with Horvath’s and Hannum’s DNAmAA [77]. In contrast to these results, Hughes and colleagues found no association between current SES and Horvath’s or Hannum’s DNAmAA in a cohort of 1,099 participants with a mean age of 58.4 [78]. The same was true for women assessed in two waves of the ALSPAC study (mean age: 28.7 and 47.4 years) and the NSHD study (mean age: 53.4 years) and Horvath DNAmAA [79]. To our knowledge, the oldest cohort analyzed in this context consisted of 490 women and men between 50 and 87 years of age (mean age: 62.2 years) [42]. This study is the only one that employs not only first-generation clocks (trained to predict chronological age) but also examines PhenoAge, a second-generation clock that aims to predict biological (phenotypic) age measures [34]. Still, no association was observed between SES (assessed as life course social class trajectory, education, and income) and DNAmAA derived from PhenoAge, Horvath’s clock or Hannum’s clock and SES in this study, either [42].

The lack of a statistically significant association between PSS and epigenetic aging in this study might be the result of several factors. First, the well-established stress marker employed here assesses perceived stress over the course of the last month before the examination. Although cortisol-mediated short-term changes in the epigenome are known, they might not be distinctive enough to translate into a detectable change in DNAmAA. It is unclear how the PSS corresponds with chronic stress in our cohort, which is often made responsible for the stress-associated adverse effects on physical and mental health [26, 27]. However, we were able to show that the PSS is associated with several relevant clinical phenotypes, such as morbidity burden, frailty, and symptoms of depression. Therefore, it seems likely that the stress marker used here does serve as a proxy of more longterm psychological stress of our participants. Second, we cannot rule out that we may have missed covariates of relevance in our regression analyses. However, this is a limitation applicable to most studies examining epigenetic markers. Furthermore, we note that we performed a detailed literature search on the topic and did not identify any additional covariates of relevance in the screened papers. Third, we cannot rule out the presence of selection/recruitment bias. The sample analyzed here is characterized by its above-average health status at baseline [53, 76]. Similarly, the average PSS scores reflected a generally low stress level, which might has impacted our results. Although we can only speculate on the reasons for these findings, a high stress level seems to be among the plausible reasons that would prevent one from voluntarily participating in a study. Finally, the lack of statistically significant findings could be the result of our sample size. However, it is unlikely that a higher number of analyzed participants would reveal clinically relevant associations as our sample size was shown to be sufficient to detect even small effect sizes in a power analysis (f^2^=0.02, power=0.8, alpha=0.05). Nevertheless, it would be of interest to repeat this analysis in an even larger sample of older participants who perceive a higher level of stress.

Strengths of this study include the usage of a well-established instrument to measure perceived stress (PSS), and the application of five different DNAm algorithms (both first- and second-generation) based on two molecular methods (MS-SNuPE and EPIC array). Despite the lack of a significant association here, further studies using individuals in a comparable age range are needed to better understand the short- and long-term consequences of acute and chronic psychological stress on biological and epigenetic age. In addition, it may be interesting to analyze biological and epigenetic age as a potential risk factor for stronger stress responses in daily life. This could help explain individual differences among participants which we observe as association between perceived stress and several health-relevant clinical outcomes.

## Conclusion

Although previous studies suggest an effect of childhood trauma on DNAmAA, the situation is less clear on the potential association between psychological stress and DNAmAA during adulthood and advanced age. In the nearly 1,000 individuals aged 64.9 years and above, we did not observe evidence for a noteworthy association between psychological stress and epigenetic aging as measured by five different epigenetic clocks.

## Data Availability

Data are available upon reasonable request. Interested investigators are invited to contact the study coordinating PI Ilja Demuth at to obtain additional information about the GendAge study and the data-sharing application form.

## Author contribution

Conceived and designed the study: VMV, DG and ID. Contributed study specific data: all authors. Analyzed the data: VMV and YS. Wrote the manuscript: VMV, JD, DG, LB and ID. All authors revised and approved the manuscript.

## References

1. Keller, A., et al., Does the perception that stress affects health matter? The association with health and mortality. Health psychology, 2012. 31(5): p. 677.

2. Dar, T., et al., Psychosocial stress and cardiovascular disease. Current treatment options in cardiovascular medicine, 2019. 21(5): p. 1–17.

3. Cohen, S., et al., Types of stressors that increase susceptibility to the common cold in healthy adults. Health Psychology, 1998. 17(3): p. 214.

4. Affleck, G., et al., A dual pathway model of daily stressor effects on rheumatoid arthritis. Annals of Behavioral Medicine, 1997. 19(2): p. 161–170.

5. Hammen, C., Stress and depression. Annu. Rev. Clin. Psychol., 2005. 1: p. 293–319.

6. Schneiderman, N., G. Ironson, and S.D. Siegel, Stress and health: psychological, behavioral, and biological determinants. Annual review of clinical psychology, 2005. 1: p. 607-628.

7. Martins de Carvalho, L., W.-Y. Chen, and A.W. Lasek, Epigenetic mechanisms underlying stress-induced depression. International Review of Neurobiology, 2020. 156: p. 87–126.

8. Fields, S.A., et al., The relationship between stress and delay discounting: a meta-analytic review. Behavioural pharmacology, 2014. 25(5 and 6): p. 434–444.

9. Sweitzer, M.M., et al., Delay discounting and smoking: Association with the Fagerström Test for Nicotine Dependence but not cigarettes smoked per day. Nicotine & Tobacco Research, 2008. 10(10): p. 1571–1575.

10. Sinha, R., Chronic stress, drug use, and vulnerability to addiction. Annals of the new York Academy of Sciences, 2008. 1141: p. 105.

11. Cohen, S., P.J. Gianaros, and S.B. Manuck, A Stage Model of Stress and Disease. Perspectives on Psychological Science, 2016. 11(4): p. 456–463.

12. Cannon, W.B., Bodily changes in pain, hunger, fear and rage.(1929.). New York, Appleton, 1953.

13. Cohen, S., D. Janicki-Deverts, and G.E. Miller, Psychological Stress and Disease. JAMA, 2007. 298(14): p. 1685–1687.

14. Chrousos, G.P. and P.W. Gold, The Concepts of Stress and Stress System Disorders: Overview of Physical and Behavioral Homeostasis. JAMA, 1992. 267(9): p. 1244–1252.

15. Cohen, S., R.C. Kessler, and L.U. Gordon, Strategies for measuring stress in studies of psychiatric and physical disorders. Measuring stress: A guide for health and social scientists, 1995: p. 3–26.

16. McEwen, B.S., Protective and damaging effects of stress mediators. New England journal of medicine, 1998. 338(3): p. 171–179.

17. Miller, G.E., S. Cohen, and A.K. Ritchey, Chronic psychological stress and the regulation of pro-inflammatory cytokines: a glucocorticoid-resistance model. Health psychology, 2002. 21(6): p. 531.

18. Dhabhar, F.S., Effects of stress on immune function: the good, the bad, and the beautiful. Immunologic Research, 2014. 58(2): p. 193–210.

19. Traustadóttir, T., P.R. Bosch, and K.S. Matt, The HPA axis response to stress in women: effects of aging and fitness. Psychoneuroendocrinology, 2005. 30(4): p. 392–402.

20. Baltes, P.B., Theoretical propositions of life-span developmental psychology: On the dynamics between growth and decline. Developmental psychology, 1987. 23(5): p. 611.

21. Charles, S.T., Strength and vulnerability integration: a model of emotional well-being across adulthood. Psychological bulletin, 2010. 136(6): p. 1068.

22. Baltes, P.B. and M.M. Baltes, Psychological perspectives on successful aging: The model of selective optimization with compensation. 1990.

23. Charles, S.T. and J.R. Piazza, Adulthood and Aging. The Corsini Encyclopedia of Psychology, 2010: p. 1–2.

24. Jylhava, J., N.L. Pedersen, and S. Hagg, Biological Age Predictors. EBioMedicine, 2017. 21: p. 29–36.

25. Horvath, S. and K. Raj, DNA methylation-based biomarkers and the epigenetic clock theory of ageing. Nature Reviews Genetics, 2018: p. 1.

26. Zannas, A.S., et al., Lifetime stress accelerates epigenetic aging in an urban, African American cohort: relevance of glucocorticoid signaling. Genome biology, 2015. 16(1): p. 1–12.

27. Zannas, A.S., et al., Correction to: Lifetime stress accelerates epigenetic aging in an urban, African American cohort: relevance of glucocorticoid signaling. Genome biology, 2018. 19(1): p. 1–1.

28. Mourtzi, N., A. Sertedaki, and E. Charmandari, Glucocorticoid Signaling and Epigenetic Alterations in Stress-Related Disorders. International Journal of Molecular Sciences, 2021. 22(11): p. 5964.

29. Zannas, A. and G. Chrousos, Epigenetic programming by stress and glucocorticoids along the human lifespan. Molecular psychiatry, 2017. 22(5): p. 640–646.

30. Gassen, N.C., et al., Life stress, glucocorticoid signaling, and the aging epigenome: Implications for aging-related diseases. Neuroscience & Biobehavioral Reviews, 2017. 74: p. 356–365.

31. Zannas, A.S., Epigenetics as a key link between psychosocial stress and aging: concepts, evidence, mechanisms. Dialogues in clinical neuroscience, 2019. 21(4): p. 389.

32. Palma-Gudiel, H., et al., Psychosocial stress and epigenetic aging, in International review of neurobiology. 2020, Elsevier. p. 107–128.

33. Felitti, V.J., et al., Relationship of Childhood Abuse and Household Dysfunction to Many of the Leading Causes of Death in Adults: The Adverse Childhood Experiences (ACE) Study. American Journal of Preventive Medicine, 1998. 14(4): p. 245–258.

34. Levine, M.E., et al., An epigenetic biomarker of aging for lifespan and healthspan. Aging (Albany NY), 2018. 10(4): p. 573.

35. McCartney, D.L., et al., Investigating the relationship between DNA methylation age acceleration and risk factors for Alzheimer’s disease. Alzheimer’s & Dementia: Diagnosis, Assessment & Disease Monitoring, 2018. 10: p. 429–437.

36. Dugué, P.-A., et al., Association of DNA methylation-based biological age with health risk factors and overall and cause-specific mortality. American journal of epidemiology, 2018. 187(3): p. 529–538.

37. Quach, A., et al., Epigenetic clock analysis of diet, exercise, education, and lifestyle factors. Aging (Albany NY), 2017. 9(2): p. 419–446.

38. Horvath, S., et al., An epigenetic clock analysis of race/ethnicity, sex, and coronary heart disease. Genome Biology, 2016. 17(1): p. 171.

39. Simons, R.L., et al., Economic hardship and biological weathering: the epigenetics of aging in a US sample of black women. Social Science & Medicine, 2016. 150: p. 192–200.

40. Ryan, J., et al., A systematic review and meta-analysis of environmental, lifestyle, and health factors associated with DNA methylation age. The Journals of Gerontology: Series A, 2020. 75(3): p. 481–494.

41. Chen, E., et al., The Great Recession and health risks in African American youth. Brain, behavior, and immunity, 2016. 53: p. 234–241.

42. McCrory, C., et al., How does socio-economic position (SEP) get biologically embedded? A comparison of allostatic load and the epigenetic clock (s). Psychoneuroendocrinology, 2019. 104: p. 64–73.

43. Boks, M.P., et al., Longitudinal changes of telomere length and epigenetic age related to traumatic stress and post-traumatic stress disorder. Psychoneuroendocrinology, 2015. 51: p. 506–512.

44. Wolf, E.J., et al., Traumatic stress and accelerated DNA methylation age: a meta-analysis. Psychoneuroendocrinology, 2018. 92: p. 123–134.

45. Han, L.K., et al., Epigenetic aging in major depressive disorder. American Journal of Psychiatry, 2018. 175(8): p. 774–782.

46. Brody, G.H., et al., Supportive family environments ameliorate the link between racial discrimination and epigenetic aging: A replication across two longitudinal cohorts. Psychological science, 2016. 27(4): p. 530–541.

47. Jovanovic, T., et al., Exposure to violence accelerates epigenetic aging in children. Scientific reports, 2017. 7(1): p. 1–7.

48. Cohen, S., T. Kamarck, and R. Mermelstein, A global measure of perceived stress. Journal of health and social behavior, 1983: p. 385–396.

49. Vetter, V.M., et al., Epigenetic clock and relative telomere length represent largely different aspects of aging in the Berlin Aging Study II (BASE-II). J Gerontol A Biol Sci Med Sci, 2018.

50. Horvath, S., DNA methylation age of human tissues and cell types. Genome Biol, 2013. 14(10): p. R115.

51. Hannum, G., et al., Genome-wide methylation profiles reveal quantitative views of human aging rates. Mol Cell, 2013. 49(2): p. 359–367.

52. Lu, A.T., et al., DNA methylation GrimAge strongly predicts lifespan and healthspan. Aging (Albany NY), 2019. 11(2): p. 303.

53. Bertram, L., et al., Cohort profile: The Berlin Aging Study II (BASE-II). Int J Epidemiol, 2014. 43(3): p. 703–12.

54. Gerstorf, D., et al., The Berlin Aging Study II–An Overview. Gerontology, 2016. 62: p. 311–315.

55. Demuth, I., et al., Cohort Profile Update: GendAge and Berlin Aging Study II (BASE-II). medRxiv, 2020.

56. Banszerus, V.L., et al., Exploring the relationship of relative telomere length and the epigenetic clock in the LipidCardio cohort. International journal of molecular sciences, 2019. 20(12): p. 3032.

57. Vidal-Bralo, L., Y. Lopez-Golan, and A. Gonzalez, Simplified Assay for Epigenetic Age Estimation in Whole Blood of Adults. Front Genet, 2016. 7: p. 126.

58. Vidal-Bralo, L., Y. Lopez-Golan, and A. Gonzalez, Corrigendum: simplified assay for epigenetic age estimation in whole blood of adults. Frontiers in genetics, 2017. 8: p. 51.

59. Vetter, V.M., et al., Seven-CpG DNA Methylation Age determined by Single Nucleotide Primer Extension and Illumina’s Infinium MethylationEPIC array provide highly comparable results. bioRxiv, 2021: p. 2021.08.13.456213.

60. Filzmoser, P., R. Maronna, and M. Werner, Outlier identification in high dimensions. Computational statistics & data analysis, 2008. 52(3): p. 1694–1711.

61. Zhou, W., P.W. Laird, and H. Shen, Comprehensive characterization, annotation and innovative use of Infinium DNA methylation BeadChip probes. Nucleic acids research, 2017. 45(4): p. e22–e22.

62. Nordlund, J., et al., Genome-wide signatures of differential DNA methylation in pediatric acute lymphoblastic leukemia. Genome biology, 2013. 14(9): p. 1–15.

63. Chen, B.H., et al., DNA methylation-based measures of biological age: meta-analysis predicting time to death. Aging (Albany NY), 2016. 8(9): p. 1844–1865.

64. Meyer, A., et al., Leukocyte telomere length is related to appendicular lean mass: cross-sectional data from the Berlin Aging Study II (BASE-II). Am J Clin Nutr, 2016. 103(1): p. 178–83.

65. Charlson, M.E., et al., A new method of classifying prognostic comorbidity in longitudinal studies: development and validation. J Chronic Dis, 1987. 40(5): p. 373–83.

66. Radloff, L.S., The CES-D Scale:A Self-Report Depression Scale for Research in the General Population. Applied Psychological Measurement, 1977. 1(3): p. 385–401.

67. Lewinsohn, P.M., et al., Center for Epidemiologic Studies Depression Scale (CES-D) as a screening instrument for depression among community-residing older adults. Psychology and aging, 1997. 12(2): p. 277.

68. Fried, L.P., et al., Frailty in older adults: evidence for a phenotype. The Journals of Gerontology Series A: Biological Sciences and Medical Sciences, 2001. 56(3): p. M146–M157.

69. Spira, D., et al., Sex-specific differences in the association of vitamin D with low lean mass and frailty–Results from the Berlin Aging Study II. Nutrition, 2018.

70. Carmel, S., Health and well-being in late life: Gender differences worldwide. Frontiers in medicine, 2019. 6: p. 218.

71. Simpkin, A.J., et al., Prenatal and early life influences on epigenetic age in children: a study of mother–offspring pairs from two cohort studies. Human molecular genetics, 2016. 25(1): p. 191–201.

72. Okbay, A., et al., Genome-wide association study identifies 74 loci associated with educational attainment. Nature, 2016. 533(7604): p. 539–42.

73. Hong, S., et al., TMEM106B and CPOX are genetic determinants of cerebrospinal fluid Alzheimer’s disease biomarker levels. Alzheimers Dement, 2021.

74. Team, R.C., R: A language and environment for statistical computing. R Foundation for Statistical Computing, Vienna, Austria. URL https://www.R-project.org, 2017.

75. Wickham, H., Elegant graphics for data analysis (ggplot2). 2009, New York, NY: Springer-Verlag.

76. Vetter, V.M., et al., Relationship between five Epigenetic Clocks, Telomere Length and Functional Capacity assessed in Older Adults: Cross-sectional and Longitudinal Analyses. medRxiv, 2021: p. 2021.10.05.21264547.

77. Fiorito, G., et al., Social adversity and epigenetic aging: a multi-cohort study on socioeconomic differences in peripheral blood DNA methylation. Scientific reports, 2017. 7(1): p. 1–12.

78. Hughes, A., et al., Socioeconomic position and DNA methylation age acceleration across the life course. American journal of epidemiology, 2018. 187(11): p. 2346–2354.

79. Lawn, R.B., et al., Psychosocial adversity and socioeconomic position during childhood and epigenetic age: analysis of two prospective cohort studies. Human molecular genetics, 2018. 27(7): p. 1301–1308.

