## Supplementary Material for "Epigenetic aging and perceived psychological stress in old age"

**Supplementary Table 1: Sex-stratified descriptive statistics of the analyzed BASE-II participants of the GendAge study (n=1,100).**

| | Women | | | | | | Men | | | | | | |
|---|---|---|---|---|---|---|---|---|---|---|---|---|---|
| | % | mean | SD | min | max | n | % | mean | SD | min | max | n | p-value |
| Chronological age (years) | | 75.72 | 3.53 | 66.41 | 94.07 | 573 | | 75.48 | 4.01 | 64.91 | 90.03 | 527 | 0.276 |
| Smoking (packyears) | | 6.30 | 13.48 | 0.00 | 114.00 | 537 | | 13.68 | 20.61 | 0.00 | 150.00 | 482 | <0.001 |
| BMI | | 26.63 | 4.68 | 17.17 | 49.68 | 573 | | 27.35 | 3.69 | 20.02 | 41.77 | 525 | 0.005 |
| Education (years) | | 14.00 | 2.91 | 7.0 | 18.0 | 527 | | 15.00 | 2.83 | 8.5 | 18.0 | 467 | <0.001 |
| Alcohol intake (yes) | 81.5 | | | | | 466 | 85.0 | | | | | 446 | 0.145 |
| Morbidity index | | 1.36 | 1.51 | 0.00 | 9.00 | 492 | | 1.43 | 1.58 | 0.00 | 9.00 | 462 | 0.478 |
| Frailty Score | | 0.79 | 0.89 | 0.00 | 4.00 | 568 | | 0.73 | 0.85 | 0.00 | 4.00 | 519 | 0.241 |
| CES-D | | 13.82 | 3.91 | 0.00 | 31.00 | 568 | | 13.25 | 3.46 | 2.00 | 35.00 | 521 | 0.010 |
| 7-CpG clock DNAmAA | | -1.02 | 6.35 | -24.37 | 25.30 | 558 | | 1.17 | 6.31 | -24.93 | 34.48 | 513 | <0.001 |
| Horvath's clock DNAmAA | | -0.43 | 3.98 | -12.31 | 23.45 | 558 | | 0.54 | 4.04 | -8.94 | 17.44 | 509 | <0.001 |
| Hannum's clock DNAmAA | | -0.72 | 3.68 | -10.80 | 12.73 | 558 | | 0.81 | 3.96 | -9.32 | 28.57 | 509 | <0.001 |
| PhenoAge DNAmAA | | -0.48 | 5.39 | -16.54 | 25.80 | 558 | | 0.62 | 5.39 | -13.51 | 20.94 | 509 | 0.001 |
| GrimAge DNAmAA | | -1.30 | 2.93 | -10.82 | 10.71 | 558 | | 1.47 | 3.27 | -8.17 | 12.85 | 509 | <0.001 |
| PSS | | 2.11 | 0.68 | 1.00 | 4.50 | 516 | | 2.04 | 0.59 | 1.00 | 4.38 | 490 | 0.082 |

Note: BMI: body mass index; CES-D Center for Epidemiologic Studies Depression Scale; PSS: Perceived Stress Scale; DNAmAA: DNA methylation age acceleration.

**Supplementary Table 2: Sex-stratified multiple linear regression analyses of morbidity index, frailty score and CES-D on Cohen's PSS in older BASE-II participants of the GendAge study.** Linear regression models were adjusted for covariates. Model 1: no adjustment; Model 2: chronological age; Model 3: Model 2 + smoking (packyears), alcohol (yes/no), BMI, and education.

| | | Women | | | | | Men | | | | |
|---|---|---|---|---|---|---|---|---|---|---|---|
| | Model | Estimate | SE | p-value | | n | Estimate | SE | p.value | | n |
| Morbidity Index | 1 | 0.268 | 0.068 | <0.001 | *** | 444 | 0.163 | 0.082 | 0.048 | * | 430 |
| | 2 | 0.264 | 0.068 | <0.001 | *** | 444 | 0.166 | 0.082 | 0.044 | * | 430 |
| | 5 | 0.310 | 0.072 | <0.001 | *** | 397 | 0.177 | 0.088 | 0.045 | * | 355 |
| Fried's Frailty Phenotype | 1 | 0.191 | 0.036 | <0.001 | *** | 511 | 0.167 | 0.042 | <0.001 | *** | 484 |
| | 2 | 0.186 | 0.036 | <0.001 | *** | 511 | 0.166 | 0.041 | <0.001 | *** | 484 |
| | 5 | 0.204 | 0.037 | <0.001 | *** | 451 | 0.154 | 0.045 | 0.001 | *** | 401 |
| CES-D | 1 | 1.109 | 0.155 | <0.001 | *** | 512 | 0.549 | 0.160 | 0.001 | *** | 485 |
| | 2 | 1.110 | 0.156 | <0.001 | *** | 512 | 0.548 | 0.161 | 0.001 | *** | 485 |
| | 5 | 1.005 | 0.161 | <0.001 | *** | 453 | 0.567 | 0.177 | 0.001 | *** | 401 |

Note: SE: standard error, CES-D: Center for Epidemiologic Studies Depression Scale.

**Supplementary Table 3: Sex-stratified multiple linear regression of Cohen's PSS on DNAmAA of five epigentic clocks and covariates.** Model 1: no covariates; Model 2: smoking (packyears), alcohol intake (yes/no), BMI, education, and genetic ancestry.

| | | Women | | | | Men | | | | |
|---|---|---|---|---|---|---|---|---|---|---|
| | Model | Estimate | SE | p-value | n | Estimate | SE | p-value | | n |
| 7-CpG DNAmAA | 1 | -0.302 | 0.264 | 0.253 | 502 | -0.196 | 0.315 | 0.534 | | 478 |
| | 2 | -0.141 | 0.293 | 0.631 | 415 | -0.153 | 0.367 | 0.678 | | 358 |
| Horvath's DNAmAA | 1 | -0.273 | 0.167 | 0.102 | 502 | -0.263 | 0.201 | 0.191 | | 474 |
| | 2 | -0.138 | 0.183 | 0.450 | 415 | -0.209 | 0.242 | 0.389 | | 356 |
| Hannum's DNAmAA | 1 | -0.165 | 0.154 | 0.286 | 502 | 0.088 | 0.196 | 0.654 | | 474 |
| | 2 | -0.042 | 0.175 | 0.811 | 415 | 0.144 | 0.233 | 0.536 | | 356 |
| PhenoAge DNAmAA | 1 | -0.148 | 0.229 | 0.518 | 502 | -0.668 | 0.265 | 0.012 | * | 474 |
| | 2 | 0.165 | 0.240 | 0.493 | 415 | -0.513 | 0.300 | 0.088 | | 356 |
| GrimAge DNAmAA | 1 | -0.007 | 0.123 | 0.953 | 502 | 0.040 | 0.162 | 0.805 | | 474 |
| | 2 | 0.066 | 0.131 | 0.611 | 415 | -0.039 | 0.177 | 0.826 | | 356 |

Note: DNAmAA: DNA methylation age acceleration; SE: Standard Error.

**Supplementary Table 4: Overview over literature of the field.**

| Study | Sample size, Study | Mean age (SD; range) | Female sex (%) | Type of stress or trauma (instrument) | Epigenetic Parameter | Main findings |
|---|---|---|---|---|---|---|
| Simons et al., 2016[6] | N=100<br>FACHS | 48.5 (9.2) | 100 | SES (income and financial pressure) Childhood trauma, Lifestyle (tobacco, alcohol intake, exercise, diet, BMI) | Hannum DNAmAA (residuals) | Association with lower income, higher financial pressure.<br>No association with childhood trauma and lifestyle. |
| Fiorito et al., 2017[8] | N=5111<br>EPIC Italy, MCCS, TILDA | 57.3 | 48.0 | SES (educational attainment, occupational position, income) | Horvath, Hannum (cell count adjusted residuals) | Metanalysis: Association between higher Horvath and Hannum DNAmAA and lower SES. |
| Hughes et al., 2018 [11] | N=1099<br>UK Household Longitudinal Study | 58.4 (14.9; 28-98) | 57.6 | SES (current income and employment, education, income and unemployment across a 12-year period, and childhood social class) | Horvath<br>Hannum | Association between low SES during childhood and higher DNAmAA. No associations between current SES and DNAmAA. |
| Lawn et al., 2018[12] | N=989 (twice) + N=773<br>ALSPAC, NSHD | 28.65 (5.54)<br>47.44 (4.42)<br>53.44 (0.26) | 100 | SES (father's occupational social class and highest current occupational social class as "high"/"low"), Psychosocial adversity during childhood | Horvath | Association between sexual abuse during childhood and higher DNAmAA. No association between SES and DNAmAA. |
| McCrory et al., 2019 [15] | N=490<br>TILDA | 62.2 (8.3; 50-87) | 50.2 | SES (social class, education and income tertiles) | Horvath<br>Hannum<br>PhenoAge | No association between DNAmAA and SES. |

References:


1. Zannas, A.S., et al., *Lifetime stress accelerates epigenetic aging in an urban, African American cohort: relevance of glucocorticoid signaling.* Genome biology, 2015. **16**(1): p. 1-12.
2. Chen, E., et al., *The Great Recession and health risks in African American youth.* Brain, behavior, and immunity, 2016. **53**: p. 234-241.
3. Simons, R.L., et al., *Economic hardship and biological weathering: the epigenetics of aging in a US sample of black women.* Social Science & Medicine, 2016. **150**: p. 192-200.
4. Fiorito, G., et al., *Social adversity and epigenetic aging: a multi-cohort study on socioeconomic differences in peripheral blood DNA methylation.* Scientific reports, 2017. **7**(1): p. 1-12.
5. Austin, M.K., et al., *Early-life socioeconomic disadvantage, not current, predicts accelerated epigenetic aging of monocytes.* Psychoneuroendocrinology, 2018. **97**: p. 131-134.
6. Hughes, A., et al., *Socioeconomic position and DNA methylation age acceleration across the life course.* American journal of epidemiology, 2018. **187**(11): p. 2346-2354.
7. Lawn, R.B., et al., *Psychosocial adversity and socioeconomic position during childhood and epigenetic age: analysis of two prospective cohort studies.* Human molecular genetics, 2018. **27**(7): p. 1301-1308.
8. McCrory, C., et al., *How does socio-economic position (SEP) get biologically embedded? A comparison of allostatic load and the epigenetic clock (s).* Psychoneuroendocrinology, 2019. **104**: p. 64-73.
